# A plasma proteomic signature links secretome of senescent monocytes to aging- and obesity-related clinical outcomes in humans

**DOI:** 10.1101/2024.08.01.24311368

**Authors:** Bradley Olinger, Reema Banarjee, Amit Dey, Dimitrios Tsitsipatis, Toshiko Tanaka, Anjana Ram, Thedoe Nyunt, Gulzar Daya, Zhongsheng Peng, Linna Cui, Julián Candia, Eleanor M. Simonsick, Myriam Gorospe, Keenan A. Walker, Luigi Ferrucci, Nathan Basisty

## Abstract

Cellular senescence increases with age and contributes to age-related declines and pathologies. We identified circulating biomarkers of senescence associated with diverse clinical traits in humans to facilitate future non-invasive assessment of individual senescence burden and efficacy testing of novel senotherapeutics. Using a novel nanoparticle-based proteomic workflow, we profiled the senescence-associated secretory phenotype (SASP) in monocytes and examined these proteins in plasma samples (N = 1060) from the Baltimore Longitudinal Study of Aging (BLSA). Machine learning models trained on monocyte SASP associated with several age-related phenotypes in a test cohort, including body fat composition, blood lipids, inflammation, and mobility-related traits, among others. Notably, a subset of SASP-based predictions, including a ‘high impact’ SASP panel that predicts age- and obesity-related clinical traits, were validated in InCHIANTI, an independent aging cohort. These results demonstrate the clinical relevance of the circulating SASP and identify relevant biomarkers of senescence that could inform future clinical studies.

## Introduction

Cellular senescence, a hallmark of aging^1^, is a state of permanent cell-cycle arrest in response to a variety of sublethal stresses, such as DNA damage, oxidative stress, metabolic imbalance, and telomere erosion^2^. Despite being in replication arrest, senescent cells remain metabolically active and secrete a plethora of proteins and other biomolecules, including cytokines, chemokines, metalloproteases, and growth factors, collectively known as the senescence-associated secretory phenotype (SASP)^3^. With increasing age, senescent cells accumulate in various tissues^1^ and, at least partially via the SASP, contribute to diverse age-related pathologies, including cognitive decline^4–7^, cardiovascular disease^8^, and immune dysfunction^9–11^, among others^12–14^. Importantly, selective elimination of senescent cells and modulation of the SASP by senotherapeutic interventions are effective strategies to improve age-associated pathologies in preclinical models^7,15,16^. Thus, developing methods to identify and eliminate senescent cells in humans is a promising goal for improving healthspan.

In recent years, senescence of immune cells, including monocytes, have been implicated as a potentially key driver of age-related pathologies. Senescence of immune cells increases with age and is thought to contribute to an age-related increase of sterile inflammation — “inflammaging” — and higher susceptibility to infectious diseases^17^. Additionally, higher levels of senescence in the immune system drive systemic aging and propagate senescence in solid organs, such as the liver, kidney and lung^18^. There is emerging evidence that senescent monocytes accumulate *in vivo* in humans. These circulating immune cells form up to 10% of the total white blood cells and are involved in pathogen recognition via Toll-like receptors (TLRs) and regulation of inflammation. A proinflammatory phenotype of monocytes has been attributed to senescence in the elderly^19^. Notably, circulating monocytes express a senescence-like signature *in vivo* in subjects with severe COVID, and SASP from these monocytes was associated with increased severity of the infection, suggesting an important role of senescent monocytes in mediating systemic inflammation in COVID patients^20^. Moreover, monocytes are promising sources of biomarkers due to their abundance in blood. Despite their potential involvement in inflammaging and biomarker potential, the role of monocyte senescence-associated proteins in aging and their potential as clinical biomarkers are poorly characterized.

Quantifying circulating biomarkers of senescence holds clinical potential for identifying outcomes tied to senescence, enabling non-invasive determination of individual senescence burden for risk stratification, and tracking the effectiveness of senotherapeutics in clinical trials^21,22^. In recent years, high-throughput proteomic studies have quantitatively profiled senescence-associated proteins in circulation and demonstrated their associations with diverse aging-related outcomes in humans^21–23^, including mortality, multimorbidity, strength and mobility. These studies leveraged human cohorts, such as InCHIANTI (Invecchiare in Chianti), BLSA (Baltimore Longitudinal Study of Aging), GESTALT (Genetic and Epigenetic Signatures of Translational Aging Laboratory Testing), Lifestyle Interventions for Elders (LIFE), and others^24–30^. However, no biomarker studies to date have comprehensively characterized the monocyte-specific SASP and evaluated its clinical utility as circulating biomarkers in humans.

The primary goal of this study is to comprehensively profile the monocyte SASP in serum-supplemented culture conditions and evaluate its clinical utility as a circulating biomarker in humans. We adopted a novel automated nanoparticle-based workflow previously leveraged for analysis of plasma^31^ that overcomes the challenges associated with mass spectrometry (MS)-based profiling of serum-supplemented medium. Thus, we completed the first comprehensive MS analysis of SASP that is not confounded by serum-free culture conditions, which are widely used for MS-based quantification of SASP from cell-culture experiments^32^. Moreover, we evaluated the senescent monocyte signatures in the plasma proteome of the BSLA study. We identified signatures of SASP, including a high-impact panel, that predict age-related and obesity-associated clinical traits in a test cohort. Remarkably, SASP-based clinical trait associations were replicated in InCHIANTI, an independent aging study.

## Results

### Optimization of senescence in monocytes and development of a biomarker discovery pipeline

To develop a rigorously validated model of cellular senescence in monocytes (**Fig. 1a**), senescent THP-1 monocyte cells were generated with multiple protocols, and an optimal method was selected, based on the expression of a combination of senescence biomarkers and cell viability. Senescence was induced in THP-1 monocytes with varying doses of gamma irradiation (IR) and assessed at various time points after IR. To test viability and proliferation, THP-1 cells in complete medium were exposed to different doses of IR (5, 7.5 and 10 Gy) and cell viability measurements at 24-h intervals up to 13 days confirmed that these doses are not lethal (**Fig. S1a**). However, cell proliferation was inhibited after exposure to 7.5 and 10 Gy as seen by reduced incorporation of 5-ethynyl-2’-deoxyuridine (Edu) in the treated cells (**Fig. 2a-b; Fig S1g**). Further, the expression levels of a panel of senescence marker mRNAs (*GDF15* mRNA, *CDKN2A* (*p16*) mRNA and *CDKN1A* (*p21*) mRNA) and mRNAs encoding SASP factors (e.g., *IL1A* and *IL6* mRNAs) were significantly higher in IR-treated cells than proliferating controls (**Fig. 2c; Fig. S1b-e**). Elevated levels of SPiDER β-gal in IR-treated cells further confirmed the stable induction of senescence 7 days after exposure to 7.5 Gy (**Fig. 2d; Fig. S1f**). Given the stable cell-cycle arrest, viability, and expression of senescence-associated mRNAs at this dose, we used 7.5 Gy IR radiation and 7 days in culture in all later experiments.

**Fig. 1.**
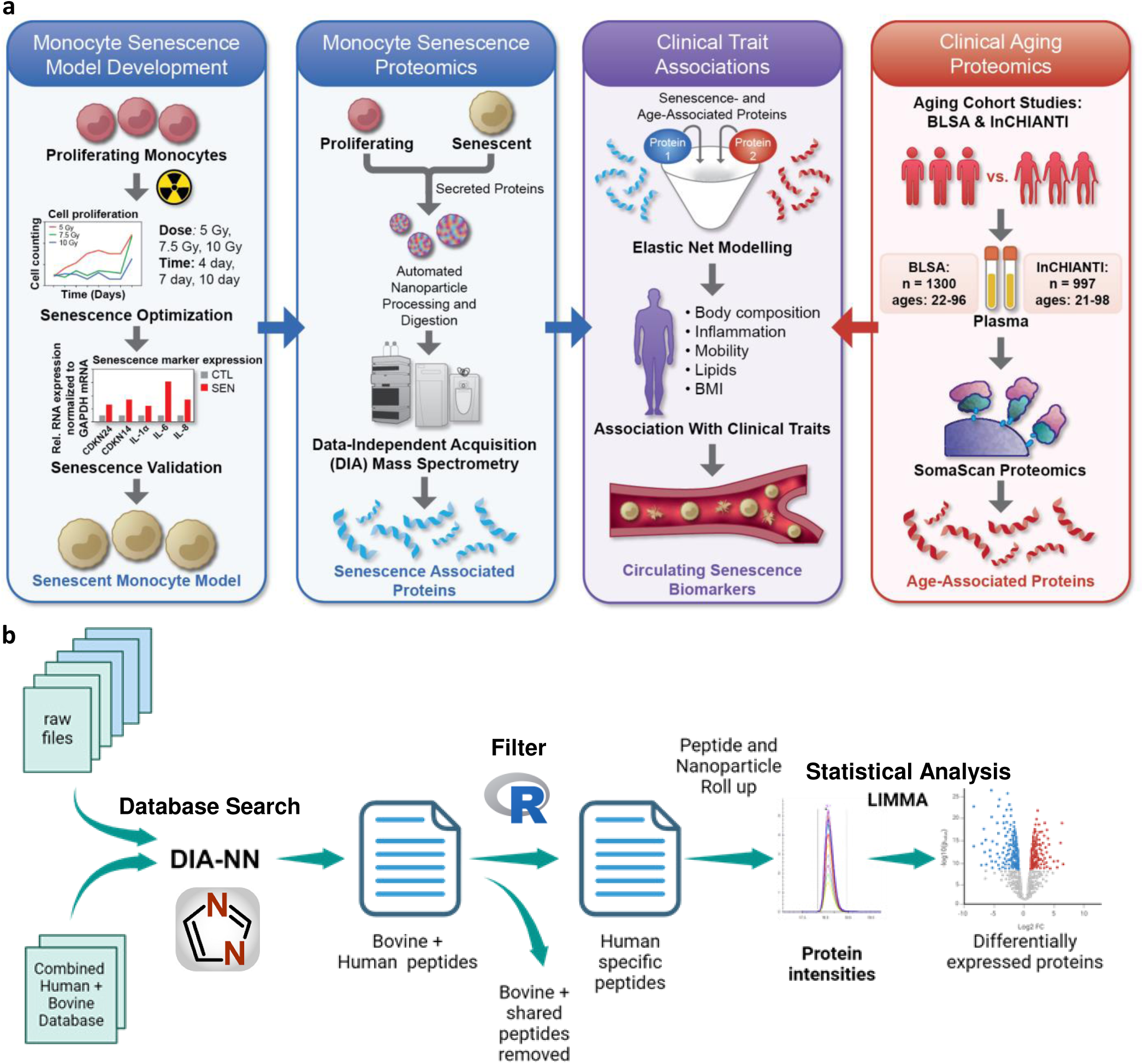
Workflow for identification of SASP signatures from the aging plasma proteome. **a,** An in vitro model of senescent monocytes was developed by exposing THP-1 cells to IR and measuring senescence markers. Further quantitative mass spectrometry proteomics was performed on the secretome of these cells using the automated nanoparticle processing and digestion platform, Proteograph. Age association of the differentially secreted proteins was evaluated using proteomic and phenotypic data from the BLSA and InCHIANTI aging cohorts. **b,** The analysis pipeline used DIA-NN to identify monocyte secretome followed by filtering out the bovine and shared peptides. Peptide quantities from each nanoparticle was rolled up to proteins to determine the differentially expressed proteins.

**Fig. 2.**
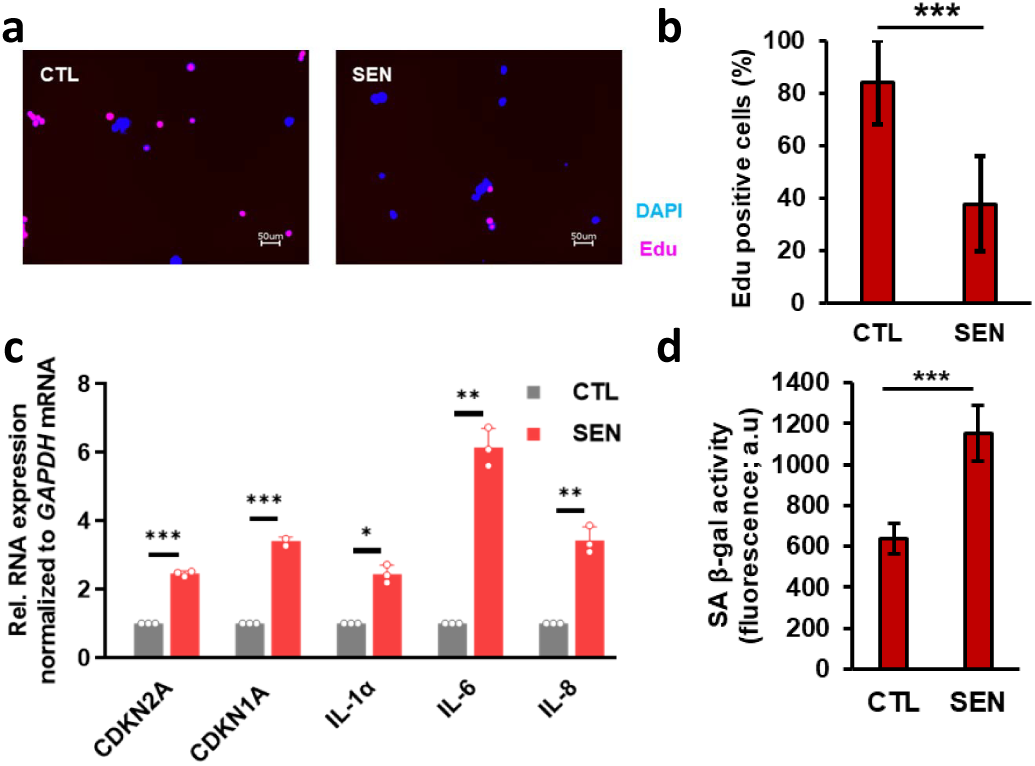
Establishing an IR-induced model of senescence in THP-1 monocytes. **a,** Representative fluorescence microscopy images from Edu incorporation assay and, **b,** corresponding quantification bar plots indicating reduced cellular incorporation of Edu by THP-1 cells 7 days after IR exposure. **c,** Bar plots showing increased expression of known senescence markers and, **d,** elevated SPiDER β-gal confirming induction of senescence in IR treated THP-1 cells. * p-value < 0.05, ** p-value < 0.01, *** p-value < 0.001.

### Identification of the monocyte SASP in standard culture conditions

MS-based proteomic analysis is notoriously challenging in serum-supplemented cell-culture conditions due to the highly dynamic range of serum proteins^33^. In standard cell-culture conditions, fetal bovine serum (FBS) introduces a high concentration of exogenous proteins that hinder the detection of endogenously secreted proteins from cultured cells at lower levels. To overcome these challenges, we applied a novel proteomic platform that was previously used to analyze blood samples^34–36^. An automated robotic platform utilizes nanoparticle-based enrichment of protein coronas for a deep and unbiased identification of proteins in high-dynamic-range samples (Proteograph^TM^ XT Assay, Redwood City, CA) ^31^. Reasoning that serum-supplemented medium faces essentially the same challenges as serum itself, we applied this instrumentation in combination with a multi-species proteomic analysis (bovine + human) to comprehensively and quantitatively profile the monocyte SASP (**Fig. 1b**). Briefly, data independent acquisition (DIA) MS-based proteomics was conducted on the secretomes of senescent and non-senescent monocytes processed with the nanoparticle workflow (n=14), as well as matched samples processed with no nanoparticle separation (n=6) to compare with a standard workflow. Using the standard workflow, 3,935 proteins (24,875 peptides) were identified, and weak separation between senescent and non-senescent secretome was evident by principal component analysis due to interference from the supplemented bovine proteins (**Fig. 3a**). In contrast, the nanoparticle-based workflow enabled the detection of 10,089 proteins (96,511 peptides) and identified differences in the two secretomes as evident in principal component analysis (**Fig. 3b-c**); these values represented a significant improvement over our reported secretome studies^30^. Additionally, even though protein expression was positively correlated on the peptide and protein level between neat and nanoparticle-processed samples (**Fig. S2a,b**), the reproducibility of protein measurements was improved after processing (**Fig. S2c**). Any peptides that matched bovine proteins, as well as those shared between human and bovine, were removed, and the remaining 6,161 human proteins were compared between senescent and non-senescent secretomes. Senescent secretomes had 3,413 increased proteins and 180 decreased proteins, compared to controls (**Fig. 3d**). Gene Ontology analysis of the top 200 upregulated proteins indicated enrichment of biological processes such as response to interferons, a known senescence-associated pathway, and transmembrane transport (**Fig. 3e**). Furthermore, a comparison of the monocyte SASP with that of fibroblasts using the SASP Atlas (http://www.saspatlas.com/)^30^ revealed 237 proteins, or 43% of the fibroblast SASP proteome, overlapped with the monocyte SASP proteome (**Fig. 3f**). Ontology analysis reveals that these proteins are involved in multiple biological processes, including cellular detoxification and regulation of apoptotic processes and oxidative stress–induced pathways (**Fig. 3g**). Interestingly, these pathways were also reported among the fibroblast core SASP^30^. Full proteomic replicate data, quantification, and statistics are available in **Table S1**.

**Fig. 3.**
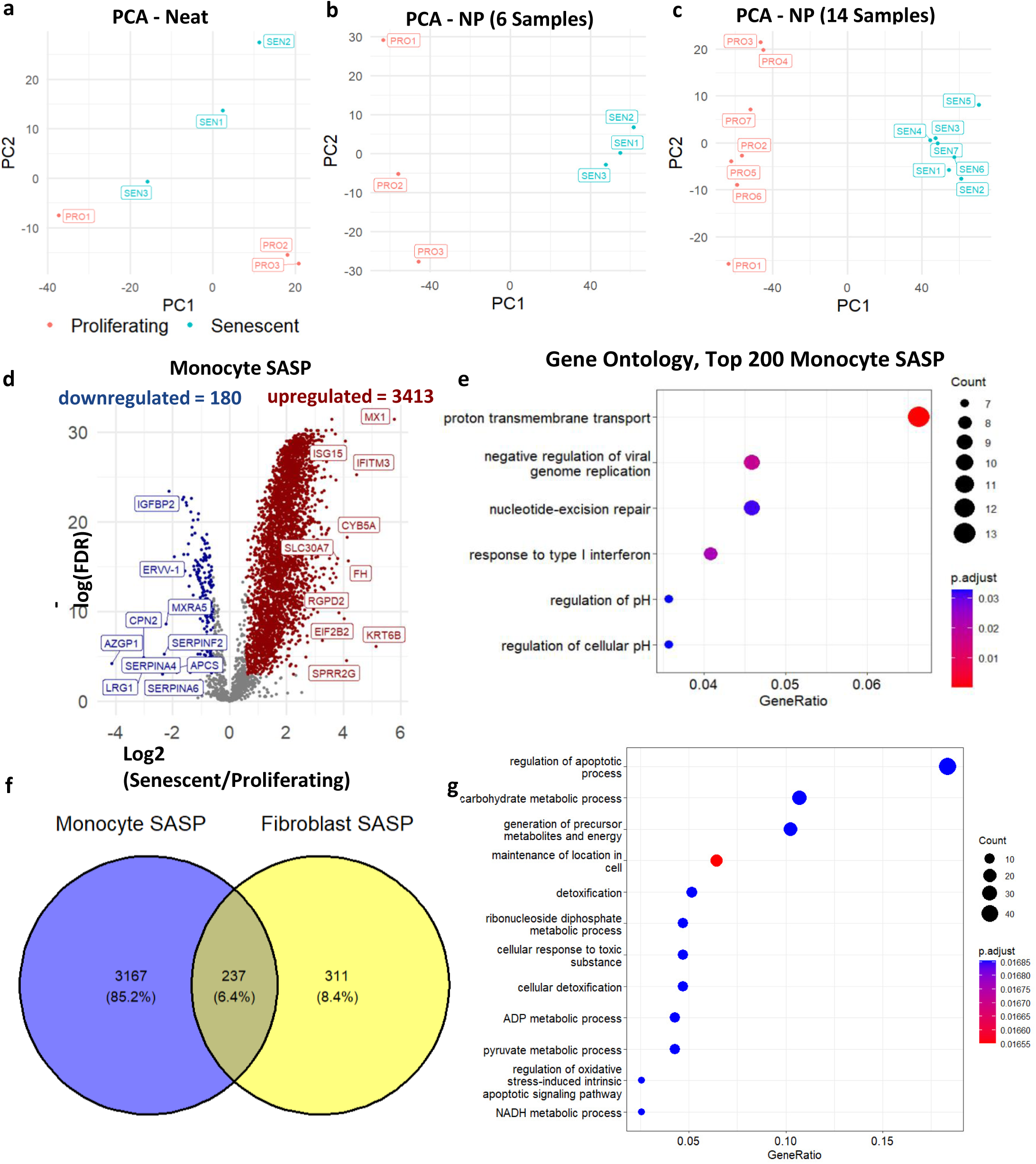
Monocyte SASP is Associated with Age in the BLSA. **a,** Principal Component Analysis (PCA) of Neat samples (protein level). **b,** PCA of 6 NP samples (protein level). **c,** PCA of 14 NP samples (protein levels). For all PCA, only proteins present in all samples were used. **d,** Monocyte SASP were identified using nanoparticle processing on the Proteograph. **e,** The top 200 differentially expressed proteins were used for ontology analysis (p-value < 0.05, q-value <0.1,Biological Process). **f,** Overlap between monocyte and fibroblast SASP. **g,** Ontology analysis of overlapping SASP proteins between monocytes and fibroblasts.

### The senescent monocyte secretome is detectable in circulating plasma in BLSA

The clinical associations of the monocyte SASP in circulation were examined in the BLSA. This longitudinal study used multiple biochemical and clinical assessments to examine the physiological and functional changes associated with healthy aging. BLSA participant characteristics are displayed in **Table 1**. This analysis utilized the plasma proteomic data from the BLSA that had been acquired using the 7K SOMAscan Assay (Somalogic Inc., Boulder, CO), which performs 7,288 protein measurements^37,38^. Among the SomaScan protein panel, 1550 monocyte SASP proteins were measured in the BLSA (**Fig. 4a, Table S2**). To evaluate the age-associated clinical relevance of the monocyte SASP, we used Spearman correlation to identify only the components of the monocyte SASP that were increasing in circulation across the lifespan in the BLSA. Of the monocyte SASP in circulating plasma, 308 proteins were upregulated and also positively associated with age (**Table S2**). To evaluate clinical traits associated with SASP regardless of covariate such as age, we utilized the full SASP panel (1550 proteins) including covariates such as age, sex, and race in downstream clinical trait associations.

**Fig. 4.**
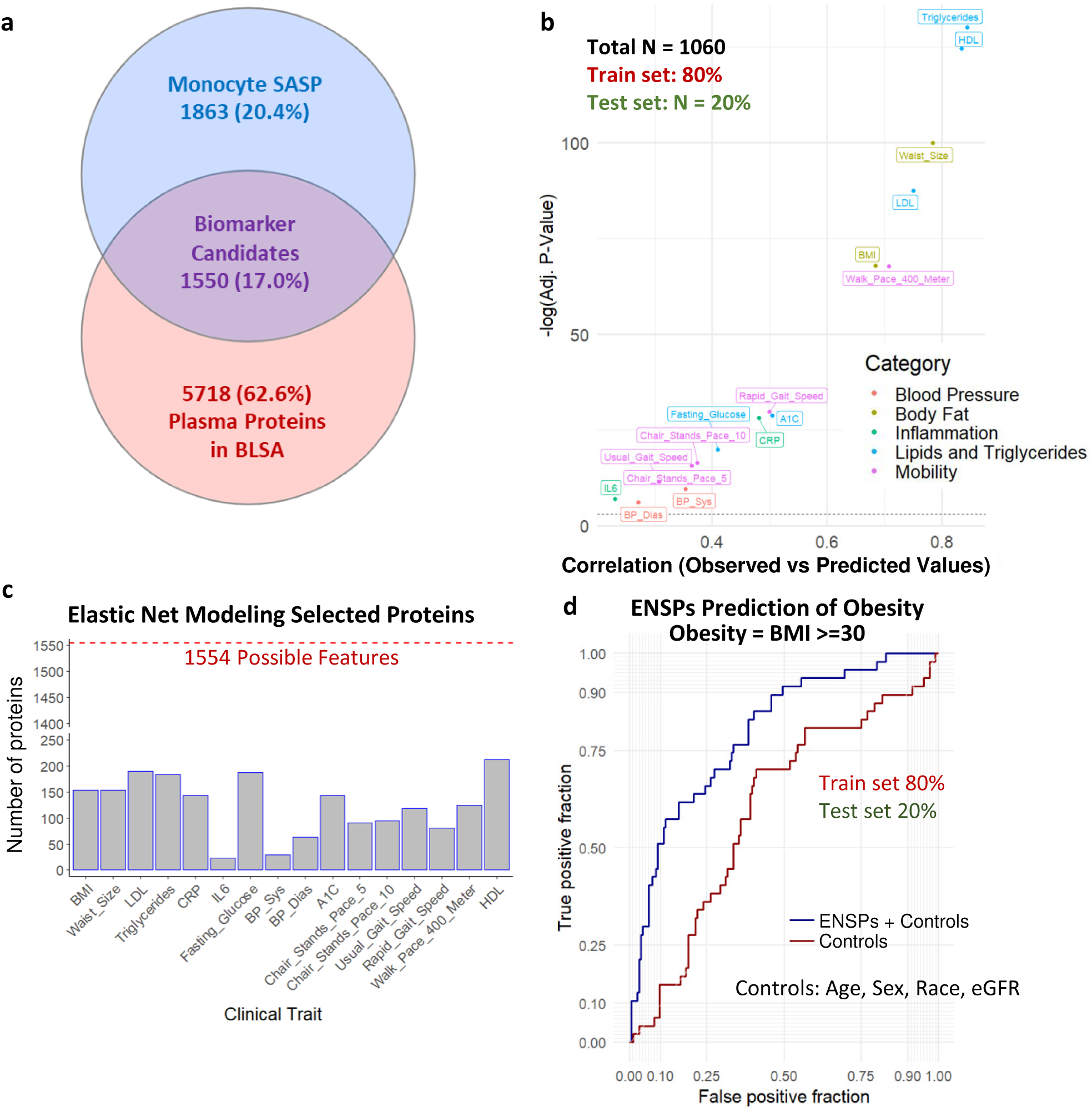
Elastic Net Modeling Using SASP of Clinical Traits in the BLSA. **a,** 1550 Monocyte SASP are detected in the BLSA 7k SomaScan. **b,** Elastic Net models were trained on 80% of the BLSA cohort and used to predict clinical traits of the remaining 20%. Spearman correlations are shown between the predicted and observed values in the test set for each clinical trait. **c,** Elastic Net modeling was used for feature selection, and the number of Elastic Net Selected Proteins (ENSPs) implicated in each clinical trait are shown. **c,** ROC plot comparing the predictive potential (80% train, 20% test) of ENSPs positively associated with BMI to predict obesity with control-only models, showing that ENSPs seem to provide additional predictive potential beyond age and other controls alone.

**Table 1.** Demographic and clinical traits in the Baltimore Longitudinal Study of Aging. Data reported as mean (standard deviation).

|  | All | 20 to 40 | 40 to 60 | 60 to 70 | 80+ | N |
| --- | --- | --- | --- | --- | --- | --- |
| Count | 1330 | 70 | 256 | 736 | 268 |  |
| age | 67.8 (14.1) | 33 (4.6) | 51.7 (6) | 70.8 (5.4) | 84 (3.2) |  |
| Female Percent | 52.3 | 54.3 | 56.6 | 51.6 | 49.3 |  |
| eGFR | 81.4 (17.7) | 106.2 (12.4) | 94.4 (13.2) | 77.1 (14.5) | 67.2 (12.8) | 1062 |
| BMI | 26.9 (4.6) | 25.1 (4.8) | 27.1 (4.8) | 27.5 (4.8) | 25.6 (3.3) | 1323 |
| Waist_Size | 88.5 (13.1) | 82.1 (12.1) | 86.2 (12.7) | 90.1 (13.5) | 88.1 (11.5) | 1283 |
| LDL | 106.4 (31.9) | 95.2 (28.1) | 111.3 (31.2) | 106.5 (33) | 103.3 (29) | 1054 |
| Triglycerides | 99.7 (50.5) | 88.6 (43.5) | 98.6 (52.6) | 102.5 (50.5) | 96.1 (49.4) | 1056 |
| CRP | 2.6 (4.4) | 1.5 (2.1) | 2 (3.1) | 2.8 (4.3) | 3.1 (6.2) | 998 |
| IL6 | 4.2 (3.5) | 3.7 (3.5) | 3.6 (2.5) | 4.3 (3.5) | 4.7 (4.6) | 996 |
| Fasting_Glucose | 96.6 (13.6) | 90.1 (6.9) | 95.1 (11.5) | 98.7 (15.4) | 95 (10.9) | 996 |
| De_Fat_Trunk | 13291.6<br>(6209.7) | 10026.7<br>(5926.6) | 13010.6<br>(6411) | 14333.8<br>(6189.1) | 11502.7<br>(5143.4) | 1022 |
| De_Fat_Body | 26497.1<br>(10343.6) | 21591.9<br>(10713.4) | 26346.4<br>(10389) | 28237.5<br>(10484.9) | 22828.3<br>(7788.7) | 1022 |
| De_Fat_Arms | 2589.9<br>(1078.8) | 2069.5<br>(1025.4) | 2582.5<br>(1110.2) | 2752.1<br>(1097.8) | 2264.5<br>(826.3) | 1022 |
| De_Fat_Legs | 9724.2<br>(4091.7) | 8729.7<br>(4340.5) | 9880.8<br>(3818.4) | 10222.8<br>(4344.1) | 8217.9<br>(2916.6) | 1022 |
| CT_Abd_SubFat | 27620.3<br>(12738.4) | 25676.1<br>(18611.6) | 27001.3<br>(12284.8) | 29057<br>(13029.9) | 23814.1<br>(9508.3) | 711 |
| CT_Abd_ViscFat | 9922.2<br>(5654.7) | 5408.9<br>(3678.7) | 7888.2<br>(4615.1) | 10761.3<br>(5847.2) | 10559.9<br>(5447.7) | 711 |
| CT_Abd_ViscSubProp | 0.4 (0.2) | 0.2 (0.1) | 0.3 (0.2) | 0.4 (0.2) | 0.5 (0.3) | 653 |
| CT_Thigh_SubFat | 8389.1<br>(4539.1) | 8094.1<br>(5255.4) | 8510.6<br>(4324.1) | 8846.6<br>(4737.3) | 6832.2<br>(3458.5) | 900 |
| CT_Thigh_IMFat | 1220.5<br>(509.8) | 895.9 (489.6) | 1174.8<br>(549.8) | 1256.4<br>(489.2) | 1306.5<br>(465.6) | 898 |
| Fat_Percent | 35.3 (9.4) | 29.5 (10.1) | 34.3 (9.3) | 37 (9.2) | 33.6 (8.5) | 1022 |
| BP_Sys | 122.9 (18.9) | 110.5 (12.6) | 118.1 (15.7) | 124.2 (18.7) | 130.4 (21.7) | 902 |
| BP_Dias | 66 (9.4) | 62.1 (7.1) | 68.1 (9.3) | 65.9 (9.6) | 64.8 (8.6) | 884 |
| A1C | 5.8 (0.6) | 5.3 (0.4) | 5.6 (0.6) | 5.9 (0.7) | 5.8 (0.3) | 965 |
| Chair_Stands_Pace_5 | 0.6 (0.2) | 0.7 (0.2) | 0.7 (0.2) | 0.5 (0.2) | 0.5 (0.2) | 975 |
| Chair_Stands_Pace_10 | 0.5 (0.2) | 0.7 (0.2) | 0.6 (0.2) | 0.5 (0.2) | 0.5 (0.1) | 972 |
| Usual_Gait_Speed | 1.2 (0.2) | 1.3 (0.2) | 1.3 (0.2) | 1.2 (0.2) | 1.1 (0.2) | 977 |
| Rapid_Gait_Speed | 1.8 (0.4) | 2.1 (0.3) | 2 (0.3) | 1.8 (0.4) | 1.6 (0.3) | 978 |
| Walk_Pace_400_Meter | 1.6 (0.3) | 1.8 (0.2) | 1.7 (0.2) | 1.6 (0.2) | 1.3 (0.2) | 929 |
| HDL | 61.4 (17.3) | 60.1 (15.5) | 60.8 (18.1) | 61.3 (16.8) | 63.1 (18.5) | 1056 |

### Monocyte SASP signatures predict mobility and obesity-associated clinical outcomes

To identify the most clinically relevant monocyte SASPs without model overfitting, elastic net modeling was performed for unbiased feature selection of proteins that associate with a diverse set of clinical outcomes, while controlling for age, sex, race, and kidney function as model covariates. Clinically relevant SASP factors were identified for a panel of clinical traits, including mobility, inflammation, body composition, and metabolism-related measurements in the BLSA (**Fig. 4b**). For each trait, a different subset and number of biologically relevant proteins were identified and subsequently referred to as elastic net-selected proteins (ENSPs) (**Fig. 4c**). One established metric for validating biomarker candidates for clinical traits is their out-of-sample predictive accuracy. This was examined in a cohort of 1,330 subjects from the BLSA, split into a training dataset (80%) and test dataset (20%). Elastic net models that include ENSPs and covariates were trained using the training dataset and used to predict clinical traits in the testing dataset. These Elastic Net models showed significant correlation between observed and predicted values (FDR < 0.05) (**Fig. 4b**) for all clinical traits, and showed highest out-of-sample trait prediction potential for Triglycerides (cor. = 0.8447), HDL (cor. = 0.8343), waist size (cor. = 0.7844), LDL (cor. = 0.7508), BMI (cor. = 0.6851), and 400-meter walking pace (cor. = 0.7085). (**Fig. 4b**), suggesting that these subsets of monocyte SASP are implicated in these clinical traits. Elastic net models also outperformed a model using covariates alone (e.g., age, sex, race and kidney function) in predicting obesity (**Fig. 4d**). After adjusting for covariates, the size of the regression coefficient for the SASP proteins in the elastic net model did not change substantially (**Fig. S3a**), suggesting that ENSPs are not simply demonstrating association with clinical traits due to their association with age and might have age-independent clinical implications.

### Association of SASP signatures with body fat depots and percentages

Because ENSPs had strikingly high predictive potential of clinical traits related to BMI and waist size, we explored whether these associations were driven by fat in specific fat depots in the body. We examined senescence markers and their predictive potential using elastic net modeling of whole-body computed tomography (CT) and dual x-ray absorptiometry scans of the BLSA participants. These measurements were used to quantify body fat content at various regions across the body, including subcutaneous fat in the limbs and abdomen, visceral fat, and intramuscular fat depots. A significant out-of-sample predictive potential was found across all body fat measures (**Fig. 5a**), utilizing ENSPs that were selected for each trait (**Fig. 5b**). Correlations between ENSPs and body fat were relatively high across different fat depots, ranging from 0.5074 to 0.7473, with the highest correlation in trunk fat and the lowest in abdominal subcutaneous fat. Moreover, total-body fat percentage was most strongly associated with senescence signatures (Spearman Cor. = 0.7913) than any individual fat depot (**Fig. 5a,d**). Because fat percentage is adjusted for overall body size, these data suggest that overall proportion of body fat, rather than the size of any specific fat depot, is associated with a monocyte senescence-associated protein signature. ENSPs were also used to predict fat percent in an independent test cohort and outperformed a covariate-only (age, sex, race, eGFR) model when predicting fat percent-based obesity (**Fig. 5c**), suggesting that ENSPs have age and covariate independent predictive potential.

**Fig. 5.**
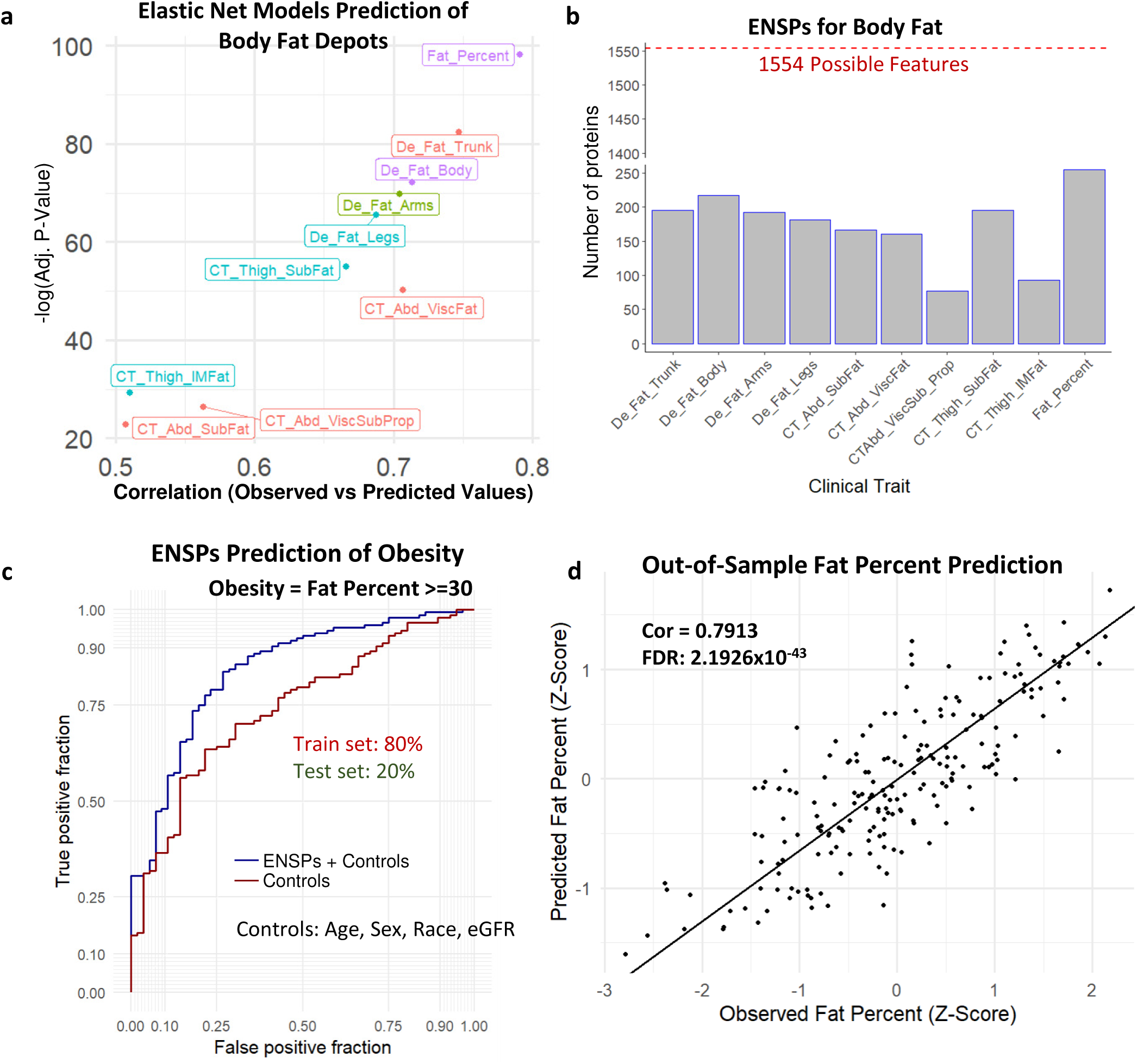
Modeling of the fat content in BLSA using SASP candidates. **a,** Elastic Net models were trained on 80% of the BLSA cohort and used to predict clinical traits of the remaining 20%. Spearman correlations are shown between the predicted and observed values of the test set for each clinical trait. **b,** Elastic Net modeling was used for feature selection, and the number of Elastic Net Selected Proteins (ENSPs) implicated in each clinical trait are shown. **c,** ROC plot comparing the predictive potential (80% train, 20% test) of ENSPs positively associated with BMI to predict obesity with control-only models, showing that ENSPs seem to provide additional predictive potential beyond age and other controls alone. **d,** The correlation between observed waist size and that predicted by Elastic Net Modeling (80% train, 20% test).

### Validation of senescence signatures in an independent aging cohort

We next sought to further validate the role of monocyte SASPs in multiple phenotypes. We performed a cross-validation analysis using plasma proteomic data from BLSA and InCHIANTI (demographics and clinical traits summarized in **Table S3**), a population-based study of older individuals living in the Chianti geographic area of Italy, which was assessed using the 1.3k SomaScan assay, an earlier version of the assay containing subset of the total protein measurements in BLSA. We identified 220 monocyte SASP candidates detected in both cohorts that were selected for further testing for clinical trait associations. ENSPs were identified for several clinical traits in both BLSA (**Fig. S4a**) and InCHIANTI (**Fig. S4b**). ENSPs identified in both cohorts were used to train linear models used for cross-validation. These linear models, when trained on the BLSA cohort, significantly predicted several clinical traits in InCHIANTI (**Fig. S4c**), and when trained on the InCHIANTI cohort, significantly predicted many clinical traits in the BLSA (**Fig. S4d**). These overlapping ENSP effectively cross-validated in both directions for several clinical traits including BMI, triglycerides, and walking pace, among others in BLSA and InCHIANTI (**Fig. 6a**), indicating a robust ability of these proteins to predict clinical traits. Additionally, a binomial model trained on overlapping ENSPs in BLSA predicted obesity in InCHIANTI better than an age and sex-only model (**Fig. 6b**). The cross-validation potential of ENSPs for a subset of clinical traits, such as BMI, blood pressure, triglycerides, and walking pace, in geographically and genetically distinct human cohorts demonstrates their robust clinical relevance.

**Fig. 6.**
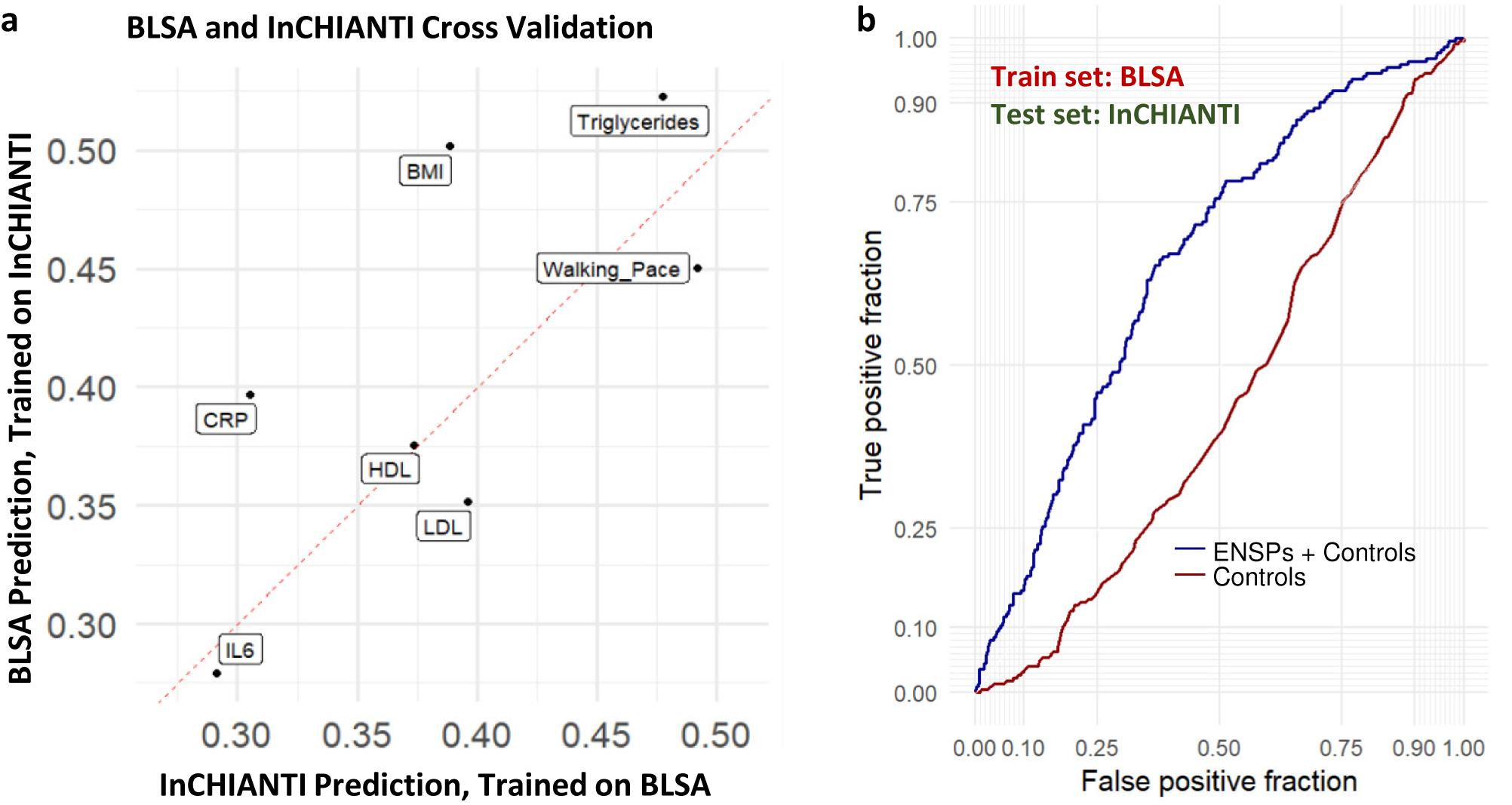
SASP-based associations show robust replication in the InCHIANTI aging study. **a,** 220 monocyte SASP were detected in both the BLSA (7k SomaScan) and InCHIANTI (1.3k SomaScan). Elastic Net modeling was used for feature selection in both Inchianti and BLSA, and linear models were constructed using only proteins selected in both studies for each trait. Spearman’s correlation of predicted values of linear models trained on the BLSA and observed values in InCHIANTI are shown on the x-axis, and Spearman’s correlation of predicted values of linear models trained on InCHIANTI and observed values in the BLSA are shown on the y-axis **b,** Binomial models were trained either using controls (age, sex) or controls + ENSPs in BLSA, then used to predict obesity in Inchianti.

### A select senescence panel robustly predicts aging- and obesity-related outcomes

To explore whether a parsimonious subset of the total SASP would capture most of the association between SASP proteins and multiple clinical traits, we prioritized and selected a smaller set of proteins based on their associations with multiple traits and relative importance. Though a different subset of SASPs was selected for each trait, consistent with previous reports^39,40^, notable features (proteins) were selected via elastic net modeling in several traits of a fourteen-trait panel. Ranking the ENSPs by the number of features in which they were implicated revealed that NQO1 and IL1RN were selected for eight of the fourteen traits, LMAN2, IGF2R, KHSRP, and CCL18 were selected for seven of the fourteen traits. Twenty-one total ENSPs were identified in at least five of the sixteen traits, seven of which were also quantified in the InCHIANTI 1.3k panel (**Fig. 7a**). Linear models of the high impact panel that were trained on 80% of the BLSA cohort significantly predicted many clinical traits of the remaining 20% test set (**Fig. 7b**). Similarly, high impact panel linear models show held-out predictive potential when trained on 80% of the InCHIANTI cohort and tested in the remaining 20% (**Fig. 7c**). Next, the high impact panel expression levels were condensed into a single continuous variable using principal component analysis. In this way, principal component 1 was used to represent each individual’s senescence burden score. Ranking the BLSA and InCHIANTI cohorts by their compositive senescence burden and plotting linear trends of the clinical traits walking pace, HDL, BMI, and CRP reveals that increasing senescence burden score shows the expected association with trait trending in the direction of poorer health (positively associated with BMI and CRP, negatively associated with walking pace and HDL), in both BLSA (**Fig. 7d**) and InCHIANTI (**Fig. S5b**). This suggests that the high impact panel could potentially be used as a proxy metric of individual senescence burden in a clinical setting.

**Fig. 7.**
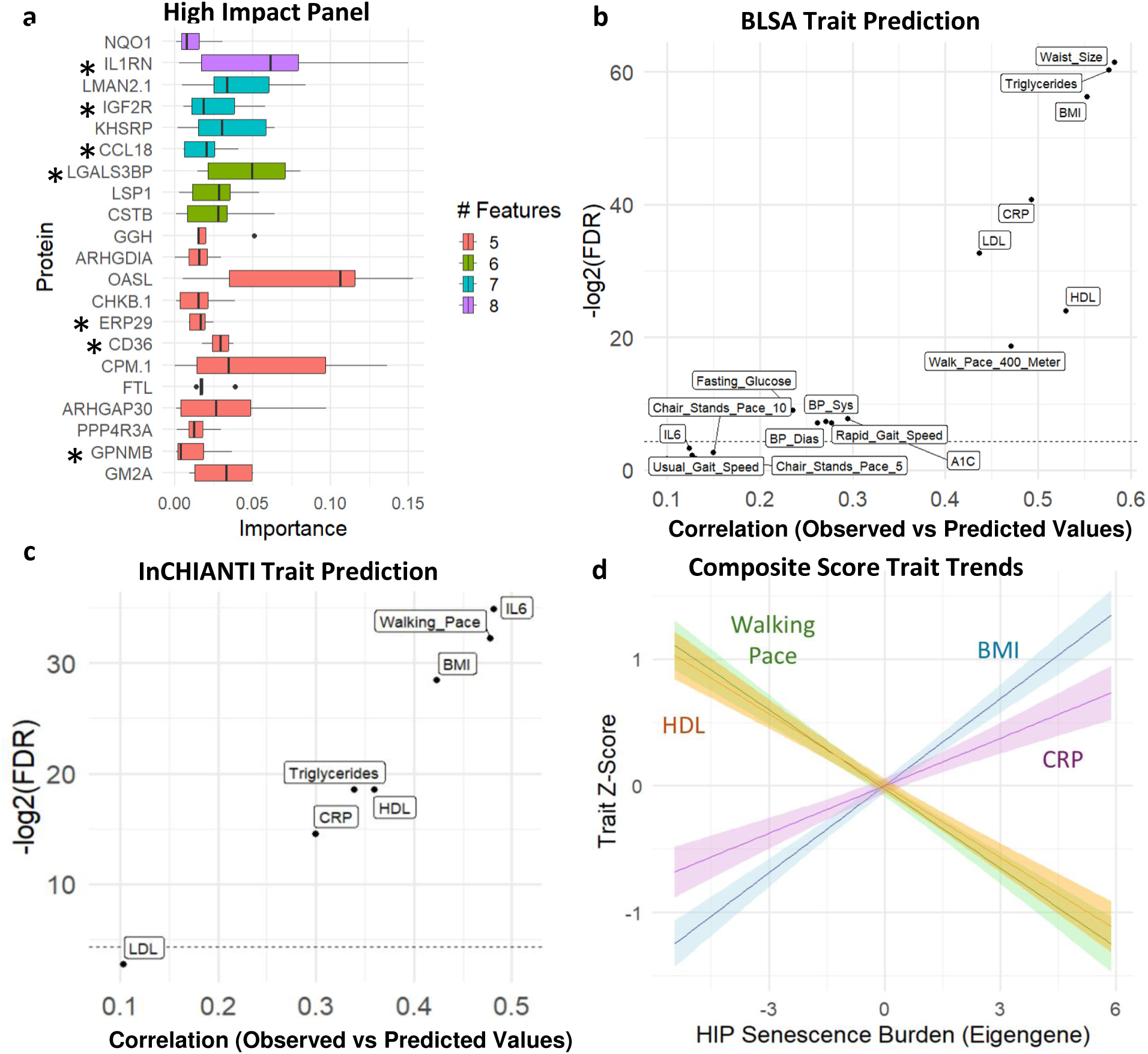
A high-impact SASP panel robustly predicts multiple clinical traits a, For a 14-trait panel, proteins were ranked by the number of features for which they were selected via Elastic Net in the BLSA, and the most frequently selected proteins are shown with their cross-trait importance on the x-axis. Only proteins that were positively associated with negative traits such as BMI and CRP, and those that were inversely associated with positive traits such as mobility were selected. Stars indicate those that were also detected in InCHIANTI. **b,** Linear models were trained on 80% of the BLSA cohort and used to predict clinical traits in the remaining 20%. Spearman’s correlation between the predicted and observed test values are shown. **c,** Linear models were trained on 80% of the InCHIANTI cohort and used to predict clinical traits in the remaining 20%. Spearman’s correlation between the predicted and observed test values are shown. **d,** Principal Component Analysis was used to condense the high-impact panel into a composite senescence burden score in the BLSA. Principal Component 1 was used to represent an eigengene for the high impact panel. With the BLSA cohort ranked from low to moderate to high senescence burden, linear trait trends reveal that positive traits HDL and Walking Pace show a negative trend, while negative traits BMI and CRP show a positive trend.

### Validation of age and covariate independent clinical relevance of senescence signatures

To evaluate the contribution of covariates such as age to the predictive potential of elastic net models, we compared covariate-only models to combined ENSP + covariate models. To select senescence signatures that are implicated in clinical traits irrespective of age and other covariates, such as race, sex, and estimated glomerular filtration rate (eGFR, a common marker of kidney function), these metrics were included in the elastic net modeling performed thus far and therefore contributed to the predictive potential of these models. To determine if the ENSPs show age and covariate independent predictive potential, analysis of variance (ANOVA) testing was used to compare covariate-only models to those that include ENSPs. ANOVA testing revealed that including ENSPs in linear models significantly improved the accuracy of covariate-only linear models (**Table S4**) Additionally, including ENSPs in covariate-only linear models substantially increased correlation coefficients (**Fig. S3a**), and ENSP-only models produced similar correlation coefficients to ENSP with covariates models (**Fig. S3a**). Due to association with body fat, a similar analysis was conducted using whole-body fat percent as an additional covariate along with age, sex, race, and eGFR to determine if ENSPs show body-fat independent predictive potential. ANOVA testing again revealed that ENSPs added significant additional predictive accuracy to models using only body fat and other covariates (**Table S4**), and adding ENSPs to body fat percent with other covariate-only linear models again increased the correlation coefficient for many clinical traits, particularly triglycerides, HDL, LDL, CRP, and fasting glucose (**Fig. S3b**). Additionally, for each clinical trait, the predictive potential of ENSPs was compared with a randomly selected group of proteins of the same group size. Permutation tests (100,000 repetitions per trait) revealed that ENSPs often demonstrated better out-of-sample trait prediction potential than randomly selected proteins (**Fig S5a, Table S5**). These results implicate senescence-associated proteins in BMI and other clinical traits independently of the effects of aging and other covariates.

## Discussion

This study applied a novel nanoparticle-based MS strategy to identify SASPs from monocytes in fully supplemented culture conditions and revealed circulating senescence signatures that predict aging-associated clinical traits in humans. MS-based proteomics allows a comprehensive unbiased characterization of the cell secretome. Yet, the large numbers of proteins, such as albumin in FBS, in most culture media are a major hurdle in MS-based analysis of SASP proteins. Our group and others addressed this issue by using serum-free media^30,32,41–44^. However, prolonged absence of serum profoundly alters cellular phenotypes, metabolism, and viability. We initially observed dramatic loss of viability and induction of differentiation under serum starvation in THP-1 cells (data not shown), necessitating a new approach compatible with serum supplementation. Serum starvation can also trigger inhibition of mTORC1, initiate autophagy^45^ and reduce protein synthesis, which greatly alter the global proteome of the cells and degrade the reliability of markers from the secretome. Moreover, mTOR is a potent regulator of the SASP in cultured cells^46,47^. Application of the automated, nanoparticle-based workflow here enabled the comprehensive profiling of the SASP in THP-1 monocytes under fully supplemented culture conditions and free of the confounders of starvation.

Our workflow adapted a recent technology that enables comprehensive proteomic analysis of the circulating proteome. This workflow leverages nanoparticles to aid in comprehensive detection of proteins in samples with a large dynamic range of protein concentrations^31,35,36,48^. In this approach, the protein mixture bound at the surface of nanoparticles in a protein sample, termed the ‘protein corona’, contains a reduced protein dynamic range and allows the detection and quantification of proteins that are normally undetectable in blood. The composition of the protein corona is reproducible and quantitative and, therefore, can be utilized for blood biomarker studies^31,36,49,50^. Because serum supplementation essentially produces the same dynamic range problem as conditioned medium, we reasoned that the same workflow would enable the comprehensive, quantitative, and unbiased profiling of the SASP under fully supplemented culture conditions. Indeed, we detected a dramatically increased number of human peptides in conditioned medium supplemented with FBS.

Despite the differences in the workflow and cell types between this study and our published SASP Atlas^30^, which focused on senescence signatures in fibroblasts, there were notable similarities in the composition of the SASPs. Here, we detected at least a threefold increase in the SASP proteins versus SASP Atlas proteins. This is likely due to a combination of factors, likely including the different MS instrumentation and the fact that SASP factor secretion is greater in serum-supplemented conditions. Nonetheless, more than 40% of the published irradiation-induced fibroblast SASP factors were also detected in the inducer-matched monocyte SASP from the current study. Among the key pathway similarities were cellular detoxification and regulation of apoptotic processes and oxidative stress–induced pathways. Furthermore, the current study identified other well-known signatures of senescent cells. Prominent among these were highly elevated interferon-related proteins (**Fig. 3e**) and, most significantly, MX1, ISG15 and IFITM3 (FDR < 1e-25). Among all SASP factors, MX1 exhibited the largest and most significant protein increase (55-fold change, FDR = 2.23e-14). Notably, multiple interferon-response associated proteins significantly elevated in the monocyte SASP (FDR < 0.05) are among the proteins increased with age in plasma of BLSA participants (FDR < 0.05), including IFI16, OAS1, IFIH1, IFNGR1, IF9, IRF4, OASL and related pro-inflammatory cytokines. Moreover, OASL was selected among other top proteins in our high impact senescence panel (**Fig 7a**) based on its association with multiple clinical traits and ranked highest in the panel in importance, based on the average magnitude of its association with each trait. Collectively, these results further reinforce the robustness of the type-1 interferon response in senescent phenotypes and highlight their potential as senescence-associated biomarkers in circulation.

Our results are consistent with the premise that senescent cells, particularly senescent monocytes, may either contribute to or be driven by declines in diverse age-related and obesity-related clinical outcomes, including loss of mobility, increased body fat (BMI, fat percentage, waist size), increased blood pressure, elevated triglycerides and lipids, elevated glucose and A1C, and inflammation (CRP and IL6). These findings are also consistent with previous studies in multiple aging cohorts identifying senescence markers that are associated with diverse clinical traits of aging, such as frailty and cognitive decline. For example, SASP proteins, such as ICAM1, MMP7 and Activin A, have been associated with a decline in physical activity in participants of the LIFE study^28^. In addition, plasma levels of 13 core SASP proteins, including CTSB, a component of the monocyte SASP, are associated with all-cause mortality and multimorbidity in the BLSA^25^. Few of the published SASP-derived protein associations were observed in the present study, highlighting the heterogeneity in senescent cell phenotypes based on cell-type. For example, GDF15 is notably missing from the monocyte SASP, while the most important SASP proteins (based on the number of associations, **Fig 7a**) identified in the present study are not among circulating senescence factors currently described^22^. These results are in line with the expected heterogeneity of senescent cells based on cell type, and that diverse SASP emerging from different senescent cell populations might drive different phenotypes. These results further suggest the importance of developing type-specific (senotype-specific) senescence biomarker signatures for drawing connections between senotype and phenotype in future studies.

One of the striking findings of this study are the robust associations between the monocyte SASP and obesity-related outcomes, such as BMI and body fat, which were among the strongest associations observed. Of note, elastic net models based on monocyte SASP predicted out-of-sample waist size, triglycerides, fat mass in multiple compartments, BMI, fasting glucose, A1C, and blood pressure. An exploratory analysis of body-fat depots in the BLSA revealed that fat deposits in different locations, including thigh, arms, abdomen, and different types, including visceral, subcutaneous, and intramuscular, were all strongly predicted by a monocyte SASP elastic net model (Spearman correlation ranging from = 0.5074 to 0.7473, **Fig 5a**). However, body fat percentage, a measure of body fat corrected to overall body size, was most strongly predicted (cor=0.7913), suggesting that fat proportion, rather than overall mass, is likely associated with senescence. The associations of monocyte SASP and body fat and related outcomes is notably independent of age and other covariates (**Fig S3a, Table S4**). These results suggest a potential link between monocyte senescence and obesity. While the direction of causality cannot be definitively determined in the present study, there is ample evidence to support obesity as a driver of cellular senescence. Culture conditions that mimic aspects of obesity, such as high free-fatty acids or glucose, can drive cells into senescence in vitro ^51–53^. Senescent cells accumulate in obesity and high-fat diet, particularly in adipose tissue^54,55^. Furthermore, the transplantation of senescent preadipocytes into mice fed high fat diet exacerbates declines in walking speed and endurance when compared with normal diet^56^. Obesity drives senescence in glial cells in mouse brains and their removal resulting in restored neurogenesis^57^. Thus, evidence strongly points to obesity as a disease-associated senescence inducer that can be decoupled from aging. Additionally, results from the CALERIE trial demonstrate the reduction of senescence biomarkers in individuals over 1-2 years of calorie restriction, further suggesting a link between senescence burden and diet, body composition and metabolism^58^. We speculate that at least some of the clinical traits described in this study can be attributed to obesity-associated senescent cells. Indeed, we observe that monocyte SASP-based elastic net models predict key obesity-associated clinical outcomes related to metabolism (fasting glucose, A1C), lipids (triglycerides, cholesterol), and blood pressure. Collectively these findings suggest the importance of considering obesity as a contributor to senescence and senescence-associated outcomes in humans. Importantly, these associations suggest that obese individuals may be among those that benefit most from senotherapeutic interventions, and we propose that this population should be considered for inclusion in future trials of senolytics and senomorphics.

Given the known age-associations of senescent cells and many SASPs, one of the potential concerns of the present study, and all studies of senescence, are the potential contribution of covariates such as aging to the associations with age-related clinical outcomes, and whether they can be separated from age-related processes. In this study, we were indeed able to show that predictive models based on SASP added value to models that include covariates (age, sex, race, and eGFR) and were clinically meaningful in predicting outcomes such as obesity. Further, to mitigate the risk of overfitting our models, which are based on large numbers of features, elastic net modeling was leveraged in this study. To further test the strength of the model, we were able to show that our elastic net model selected on monocyte SASP far outperformed linear models based using the same number of randomly selected proteins across the 7k SomaScan assay (**Fig S5a**). To ensure the robustness of the findings, we report associations are based on prediction out-of-sample clinical outcomes (independent of the training set), including a subset of the clinical associations that replicated across BLSA and InCHIANTI (**Fig 6a**).

One of the clinically meaningful findings of his study was that, despite the large number of total SASP proteins identified, a relatively small panel of these also robustly predicted a set of age- and obesity-associated clinical outcomes, including inflammation (IL6, CRP), lipids (HDL, LDL), glucose (A1c, fasting glucose), blood pressure, walking speed and pace, and BMI. Notably, even though a fraction of this panel was measured in both BLSA and InCHIANTI for replication, multiple predictions were replicated across aging studies, supporting the robustness of using selected high impact proteins, for clinical associations. A defined panel of proteins may be clinically advantageous in that the full panel, or selected proteins, can be more readily tested and applied in multiple studies without building new models or making costly measurements of large numbers of proteins. In future studies, it will be of interest to further validate this panel in diverse human cohorts and test their utility to predict a range of aging- and obesity-related outcomes. Several proteins in the panel are consistent with senescence biology and have known associations with outcomes with elevated senescent cell burden. Notably, the cytokine CCL18 (also known as PARC) previously showed the strongest association with mortality among 28 SASPs in a study of 1923 individuals over the age of 65^59^, and has been associated with disease-progression or negative outcomes in a range of diseases including cancer ^60^, atherosclerosis ^61^, and lung disease ^62^. Glycoprotein nonmetastatic melanoma protein B (GPNMB) is a senescence-associated protein that, when targeted with a senolytic vaccine, results in the reduction of senescence burden and improvements in aging and obesity-related outcomes in mice, including improved glucose homeostasis on high fat diet, and reduced aortic plaque size in APOE KO mice^63^. LGALS3BP is a previously reported core SASP^30^, is associated with diverse malignancies^64^ and sepsis^65^. In future studies, it will be valuable to validate the associations of the set of proteins in the high impact senescence panel with elevated senescence burden and evaluate their potential roles as either drivers or biomarkers of disease outcomes.

This study has several limitations. SASP factors in plasma can be contributed by a variety of cells and tissues in the body. Thus, it is not possible to track the originating tissues of SASPs in circulation or to verify whether circulating proteins were released by senescent cells or other secreting cells with common secretory factors, such as activated immune cells. Senescence signatures are numerous and heterogeneous by cell type^3^, and examining clinical associations of senescence signatures from a variety of tissue types is warranted. Studying the SASP from specific cell types can help dissect the role of individual cells in the progression of age-associated clinical traits. In future studies, it may be of interest to identify tissue-specific (senotype-specific) senescent signatures and examine their clinical associations in human cohorts. These studies may shed light on the contributing tissue and senotype-specific senescent cell populations on aging- and obesity-related outcomes and identify more sensitive and specific biomarkers. One limitation of the cross-study validation performed is the difference in the proteomic panels applied in each study, where the BLSA was performed on the newer generation of the SomaScan panel versus the InCHIANTI study. While this affected the strength of the predictions, multiple outcomes remained significant with smaller panels, highlighting some of the more robust associations. It will be valuable going forward to test the cross-study validations both in studies that utilize the complete proteomic assay, and in studies that utilize different proteomic platforms such as UK Biobank, to better understand the predictors that are most robust to differences in proteomic methods. Finally, longitudinal proteomic measurements will be useful in future studies for evaluating whether SASP protein trajectories can be more sensitive in predicting clinical outcomes.

In summary, we showed that SASP factors from monocytes have a high association with aging-associated clinical traits and can serve as biomarkers to predict biological aging. Using nanoparticle-based enrichment coupled with MS enabled comprehensive characterization of the secretome from senescent monocytes in culture in serum-supplemented culture conditions. Our results highlight a novel approach to study the cellular secretome under physiological conditions. Moreover, this study sheds light on clinical associations of circulating monocyte SASPs in human longitudinal studies and identifies possible biomarkers of senescence that could potentially inform future senotherapeutic trials in obese and aged individuals.

## Methods

### Cell culture and senescence induction

THP-1 human monocytes (ATCC, Manassas, VA; #TIB-202) were cultured in RPMI 1640 medium (Thermo Fisher Scientific Inc., Waltham, MA; #11875119) supplemented with 10% FBS (Thermo Fisher Scientific; #26140079) and 1% pen strep (Thermo Fisher Scientific; # 15070063) in a 20% O_2_, 5% CO_2_ incubator. To determine the optimum conditions for induction of senescence in monocytes, proliferating THP-1 cells were exposed to different doses of ionizing (γ) radiation (IR; 5, 7.5, and 10 Gy) using the Gammacell 40 Exactor, a Cesium-137 based irradiator (Nordion Inc., Canada); thereafter, cell viability and senescence markers were assessed over different timepoints. Cell viability was measured using the CellTiter-Glo® Luminescent Cell Viability Assay (Promega Corporation, Madison, WI; #G7570) as per the manufacturer’s instructions. IR radiation of 7.5 Gy and culture up to 7 days were optimum for senescence induction and were used for all further experiments. Proliferating cells were used as non-senescent controls for all experiments.

### EdU incorporation assay

Cell proliferation was assessed using the Click-iT™ Plus EdU Cell Proliferation Kit (Thermo Fisher Scientific; #C10640), following the manufacturer’s instructions. Briefly, 20,000 cells per well were seeded in 100 μL of medium in 96-well plates. At 6 days after irradiation, cells were incubated with 20 μM 5-ethynyl-2’-deoxyuridine (EdU) overnight at 37°C. Next day, cells were fixed using 3.7% formaldehyde for 15 min, followed by permeabilization with 0.5% Triton X-100 for 20 min. Cells were then incubated with Alexa Fluor picolyl azide and Hoechst 33342 dye for 30 min each. The resulting images were captured by a fluorescence microscope (BZ-X Analyzer, Keyence Corporation, Itasca, IL). The percentage of cells that showed EdU incorporation corresponded to the percentage proliferating cells.

### Quantitative PCR (qPCR) analysis

Total RNA was isolated using a Direct-zol RNA Miniprep Kit (Zymo Research, Irvine, CA; #R2052), and 500 ng of total RNA was reverse transcribed using Maxima reverse transcriptase (Thermo Fisher Scientific; # EP0741) and random hexamers. For reverse transcription followed by quantitative RT-qPCR analysis, 0.1 µl cDNA was employed with 250 nM of gene-specific primers (**Table S6**) and KAPA SYBR® FAST qPCR Kits (KAPA Biosystems Inc., Wilmington, MA; #) on the QuantStudio 3 Realtime PCR system (Thermo Fisher Scientific). Relative RNA levels were calculated after normalizing to *ACTB* mRNA encoding the housekeeping protein β-actin using the 2^−ΔΔCt^ method. QPCR experiments were performed with seven biological replicates of IR-treated and proliferating controls. Statistical analysis was performed using GraphPad Prism v10. Results are presented as the mean ± SD. Comparisons between the two groups were made using students *t*-test. Statistical significance was considered at p < 0.05.

### Senescence-associated β-galactosidase (SA-β-gal) assay

Senescence-associated β-galactosidase (SA-β-gal) activity was assessed using the Cellular Senescence Plate Assay Kit - SPiDER-βGal (Dojindo Molecular Technologies Inc., Rockville, MD; #SG05), following the manufacturer’s instructions. Briefly, 150,000 cells were seeded in 2 mL of medium in 6-well plates. On the 7^th^ day after IR, cells were lysed and incubated with SPiDER β-gal for 1 h at 37°C, and SPiDER β-gal fluorescence intensity was measured at 535 nm excitation and 580 nm emission using a microplate reader.

### Sample preparation for mass spectrometry

SASPs from senescent and proliferating THP-1 monocytes were collected, based on a modified version of the protocol from Neri et al.^32^, adjusting the protocol so that complete medium (supplemented with 10% FBS) was used in place of serum-free medium because of the downstream processing of samples with the Proteograph workflow. Briefly, THP-1 cells were grown in T75 flasks to sub-confluence prior to induction of senescence. For senescent samples, cells were shifted to fresh complete medium 7 days after irradiation, and conditioned medium containing SASP was collected after 48 h. Conditioned media from proliferating cells were similarly collected after 48-h incubation in fresh complete medium. Conditioned medium was then centrifuged at 10,000 x g for 15 min to pellet down cell debris and concentrated using 3 kDa molecular mass cut-off filters. One milliliter of each conditioned medium sample was then aliquoted into tubes for downstream preparation, for a total of 14 samples (seven senescent and seven non-senescent).

The conditioned medium samples were processed as described^34^ on the SP100 Automation Instrument coupled with Proteograph XT Assay Kits (Seer, Inc). Briefly, samples were incubated with two chemically distinct nanoparticle suspensions supplied in the assay kits. Nanoparticle-bound proteins were captured by incubation and a series of gentle washes. The resulting protein coronas were reduced, alkylated, and digested with Trypsin/Lys-C to generate peptides for MS analysis. The peptide mixture was desalted using solid phase-extraction. All steps in the preparation of peptides were conducted automatically by the SP100, which produces two peptide fractions per sample, one fraction for each nanoparticle suspension.

In addition to the preparation of 14 nanoparticle-processed samples, six matched neat samples were prepared (three senescent and three non-senescent). Neat samples were processed in parallel with the other samples, except no nanoparticles were added and no protein corona was captured. Essentially, neat samples are equivalent to a standard digestion protocol and are used for comparison against standard methods. After processing of samples with the Proteograph XT workflow, 34 peptide samples were generated for downstream MS analysis: 28 experimental samples (14 replicates with two nanoparticle fractions each) and six matched neat samples.

### Liquid chromatography**–**mass spectrometry

All samples were analyzed using a Vanquish Neo UHPLC system coupled to an Orbitrap Astral mass spectrometer (Thermo Fisher Scientific) with a NanoSpray Flex source (Thermo Fisher Scientific). Peptides (400 ng of each sample) were loaded on Acclaim PepMap 100 C18 (0.3 mm ID x 5 mm) trap column and then separated on a 50-cm μPAC analytical column (PharmaFluidics, Belgium) at a flow rate of 1.0 µL/min using a gradient of 4–35% solvent B (0.1% formic acid in acetonitrile) mixed into solvent A (0.1% formic acid in water) over 20.8 min and a total run time of 24 min. The MS data were acquired in data-independent acquisition (DIA) mode with a normalized HCD-collision energy of 25% and a default charge state of +2. MS1 spectra were acquired in the Orbitrap every 0.6 s at a resolving power of 240,000 at m/z 200 over m/z 380–980. The MS^1^ normalized AGC target was 500% (5×10^6^ charges) with a MaxIT of 5 ms. For MS/MS experiments, the DIA experiment was set to have a 3 m/z isolation window making 199 DIA scan events across the precursor isolation windows spanned 380–980. MS2 scans were collected from 150-2000 m/z. DIA MS^2^ scans were acquired in Astral analyzer with a normalized AGC target of 500% (5×10^4^ charges) to better control the ion population using MaxIT that was set to 5 ms. Window placement optimization was turned on. A source voltage of 1500 V and an ion transfer tube temperature of 280 °C were used for all experiments.

### Protein identification and quantitative analysis pipeline

Data were analyzed using the Proteograph^TM^ Analysis Suite (PAS) and the DIA-NN v1.8.1 algorithm to perform the peptide identification and quantification using an *in silico*-predicted library, based on a combined database containing both Uniprot human and bovine proteomes (4/2022 builds, 105,533 entries including isoform and TrEMBL). Match between runs was enabled. Otherwise, all DIA-NN settings were set to default. FDR was set to 1% for filtering identifications at the peptide and protein group levels. Peptide quantification was performed using a max representation approach, where the single quantification value for a particular peptide represents the quantitation value of the nanoparticle most frequently measured across all samples, and standard peptide-to-protein rollup was used on selected peptide values. Furthermore, to specifically identify the monocyte SASP and eliminate proteins originating from the FBS and unambiguously remove non-human peptides, all the peptides that mapped to the bovine proteome, or both human and bovine proteins, were removed from further analysis. Scaled intensities of human-unique peptides were aggregated to generate protein intensities using the MaxLFQ algorithm, implemented in the R/Bioconductor package iq (version 1.9.12) ^66^. Differential analysis was conducted on the protein intensities between PRO and SEN samples using the R/Bioconductor package limma (version 3.58.1) ^67^. Multiple testing was corrected using the Benjamini-Hochberg procedure.

### Pathway analysis

Gene Ontology analysis was performed using ClusterProfiler version 4.6.0 ^68,69^ in Rstudio to identify the biological processes and molecular functions enriched among the differentially expressed proteins. A background list of all proteins was utilized in each analysis. In Fig. 3e, the background list of proteins included all human proteins identified by MS. In Fig. 3g, the background list consisted of all proteins identified in both the fibroblast and monocyte secretomes. In Fig. 4c, the background list of proteins included all proteins that overlapped between the SomaScan 7K assay and the monocyte secretome proteins. All pathways depicted in main figures were statistically significant after background correction and multiple-testing correction, with adjusted p < 0.05, calculated by the Benjamini-Hochberg Procedure. Analysis was conducted using R version 4.2.0 (R Development Core Team, Vienna, Austria) and RStudio 2023.06.0-421 (RStudio, Boston, MA).

### Clinical data

We used clinical data from the BLSA and InCHIANTI cohorts to determine the association of senescence markers with physiological markers of health and aging. The BLSA study is a population-based study that started in 1958 to evaluate the contributors of healthy aging in subjects recruited from the DC/Baltimore metropolitan area that are 20 years old and older ^70–72^. It involves a data collection of clinical parameters, such as waist circumference, BMI and blood pressure that are assessed during a standard medical exam. Blood tests were performed at a Clinical Laboratory Improvement Amendments certified clinical laboratory at Medstar Harbor Hospital, Baltimore, MD. Total cholesterol was measured using alkaline phosphatase, HDL and LDL with dextran magnetic beads, triglycerides with colorimetric methods, glucose with glucose oxidase using the Vitros system (Ortho Clinical Diagnostics, Raritan, NJ). Serum inflammatory markers IL6 (R&D System, Minneapolis, MN) and CRP (Alpco, Salem, NH) were measured with enzyme-linked immunosorbent assay (ELISA). HbA1C levels were measured using liquid chromatography by an automated DiaSTAT analyzer (Bio-Rad, Oakland, CA). Grip strength was measured three times on each of the right and left hand and the highest average grip strength was reported. Usual gait speed was measured in two trials of a 6-m walk and the faster time between the two trials is used for analysis. Body fat measurements were made using CT/dual x-ray absorptiometry scans. The BLSA protocol (03AG0325) was approved by the institutional review board of the National Institute of Environmental Health Science, part of the National Institutes of Health. To avoid possible confounding effects of medication use, patients taking diabetes or hypertension medication had their A1C or blood pressure values excluded from statistical analysis.

The InCHIANTI is a similar population-based study of aging conducted in the Chianti region of Tuscany, Italy previously described in more detail ^73^. Residents from the population registry of Greve in Chianti (a rural area) and Bagno a Ripoli (near Florence) ranging in age from 21 to 102 years participated in the study. The study (exemption #11976) protocol was approved by Medstar Research Institute (Baltimore, Maryland), the Italian National Institute of Research and Care of Aging Institutional Review, and the Internal Review Board of the National Institute for Environmental Health Sciences (NIEHS). All participants provided written informed consent.

### SOMAscan assay

The BLSA plasma proteomic study involved profiling for 7,596 SOMAmers using the 7K SOMAscan Assay, whereas the InCHIANTI involved profiling for 1322 SOMAmers using the 1.3K SOMAscan Assay at the Trans-NIH Center for Human Immunology and Autoimmunity, and Inflammation (CHI), National Institute of Allergy and Infectious Disease, National Institutes of Health (Bethesda, MD). Proteomic assessment and data normalization for each of the assays were conducted as reported^37,74^. Plasma protein concentrations were directly proportional to the abundance of the SOMAmer reagents that were reported in relative fluorescence units. The reliability and variability of the SomaScan assay measurements has been previously rigorously evaluated and applied in the BLSA ^37,75^.

### Elastic-net modeling

To avoid overfitting of simple linear models, Elastic-net modeling, a penalized linear regression machine learning technique, was used for feature selection of the most biologically relevant proteins. Elastic-net modeling was used due to its ability to perform unbiased feature selection, and because it can cope with high collinearity of features. Elastic Net contains two tuning parameters, alpha and lambda, that determine the nature of feature selection used. The value for alpha can range from 0 (Ridge regression with no feature elimination) to 1 (Lasso regression which selects the smallest number of features). An alpha of 1 was used to apply the strictest level of feature selection and reveal the smallest number of highly relevant features. The lambda parameter determines the strength of the penalty, with a higher penalty eliminating more features. The package glmnet^76^ (version 4.1-6) randomly selected subsets of the dataset for cross-validation to find the lambda value with the smallest mean squared error. Due to its feature tuning method, this package produces slightly variable results. To account for this and to reduce variability, the package was run 100 times for each trait, and the most accurate penalty term was found across all runs.

## Data Availability

Data Availability: All raw mass spectrometry data files and associated quantitative and statistical reports, metadata, and supplemental data are available on MassIVE (dataset identifier: MSV000095315). FTP download link: ftp://massive.ucsd.edu/v08/MSV000095315/.

Code Availability: R scripts for the core elastic net analysis described are available at https://github.com/geroproteomics/EN_Repeat/blob/main/EN_Repeat.

Other data are available upon reasonable request to the authors.

ftp://massive.ucsd.edu/v08/MSV000095315/

https://github.com/geroproteomics/EN_Repeat/blob/main/EN_Repeat.

## Data Availability

All raw mass spectrometry data files and associated quantitative and statistical reports, metadata, and supplemental data are available on MassIVE (dataset identifier: MSV000095315). FTP download link: ftp://massive.ucsd.edu/v08/MSV000095315/.

## Code Availability

R scripts for the core elastic net analysis described are available at https://github.com/geroproteomics/EN_Repeat/blob/main/EN_Repeat.

## Acknowledgements

This work was supported by the National Institute on Aging (NIA) Intramural Research Program (IRP), NIH. N.B. was supported by a SenNet NIH Common Fund Grant (NIA U54 AG079779, PI: Elisseeff) and a Hevolution GRO grant (HF-GRO-23-1199068-44). We gratefully acknowledge Lauren Brick and NIDA/NIA Visual Media for assistance in figure preparation and Gary Howard for editing of the manuscript.

