## Supplemental Figures for "A plasma proteomic signature links secretome of senescent monocytes to aging- and obesity-related clinical outcomes in humans"

### Supplemental Figure 1

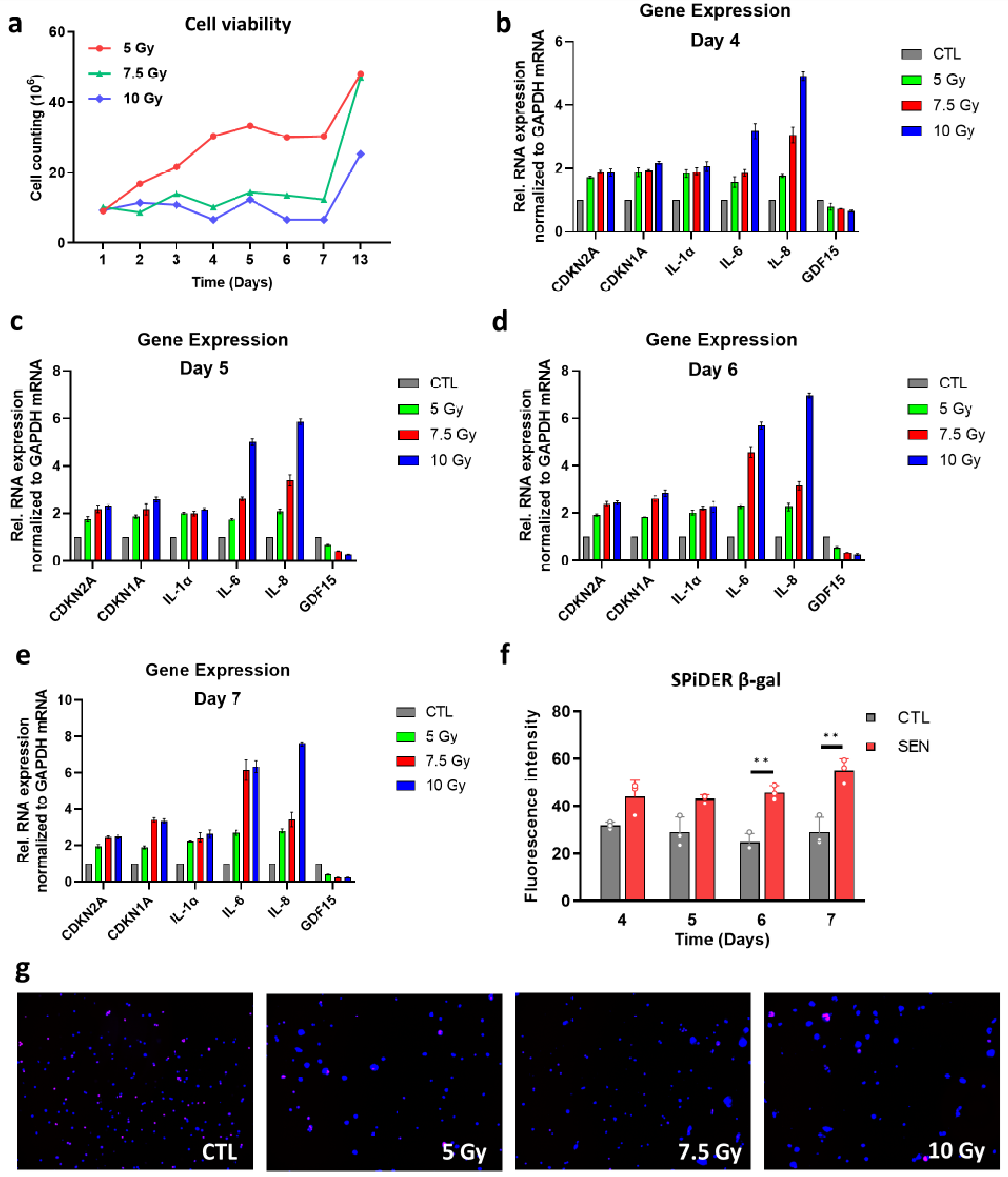


**Fig S1. Optimization of IR induced senescence in THP-1 monocytes. a,** Bar plot indicating the cell number after exposure to different doses of IR. **b, c, d,** and **e,** Bar plots showing increased expression of known senescence markers over time number after exposure to different doses of IR. **f,** Bar plots showing elevated SPiDER β-gal confirming induction of senescence in IR treated THP-1 cells. **g,** Representative fluorescence microscopy images from Edu incorporation assay after exposure to different doses of IR.

### Supplemental Figure 2

**A**

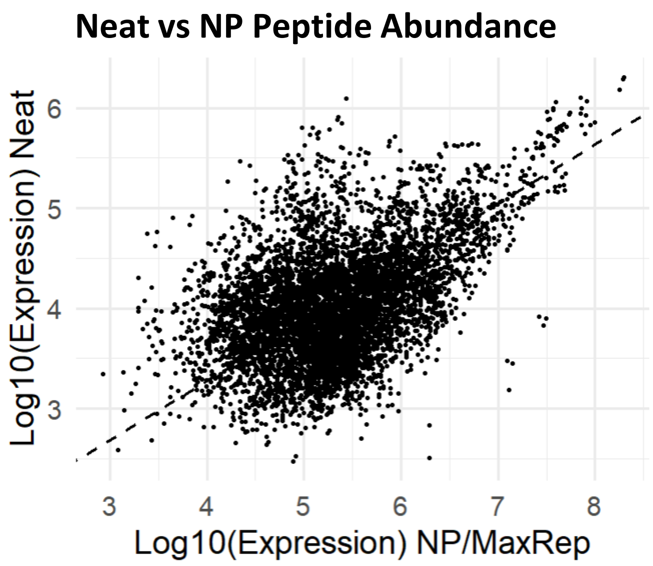


Cor: 0.40098
P-Value: 8.826397e^-243

**B**

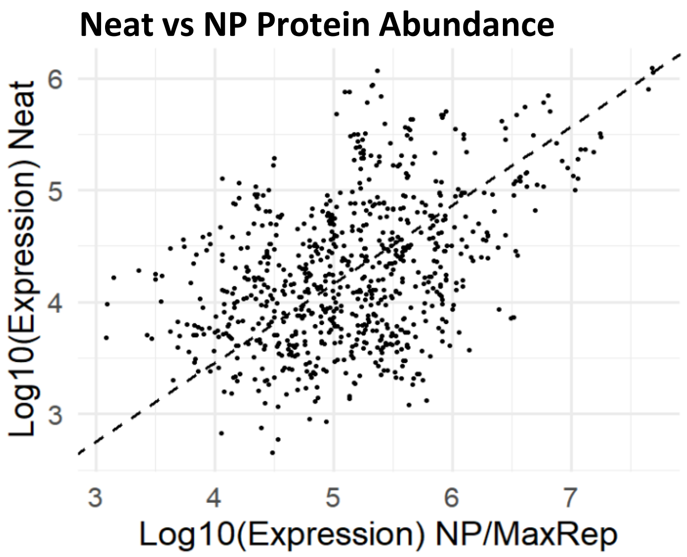


Cor: 0.36775
P-Value: 4.55432e^-26

**C Comparison of Neat and NP Protein CV**

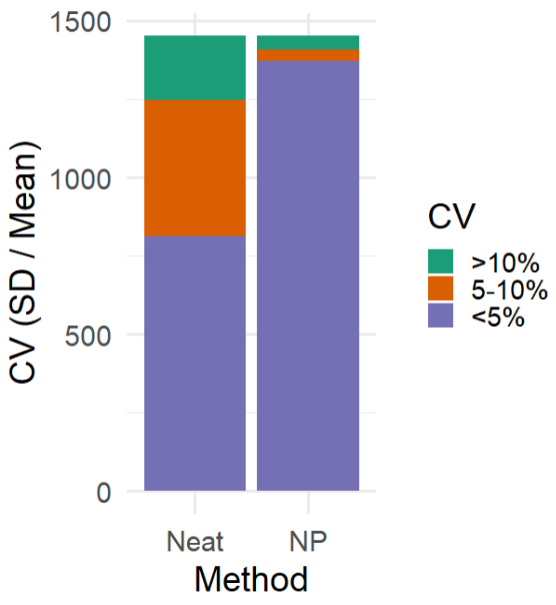


**Fig S2. Comparison of nanoparticle and neat proteomic workflows. a,** Peptide expression level is compared between quantification methods of 3 proliferating and 3 senescent monocyte samples, using NanoParticles or Neat (control). Only peptides present in at least 4 of 6 samples, showing differential expression (Pval < 0.05, t-tests) in neat samples were included (n = 1,149). **b,** Protein expression level is compared between quantification methods of 3 proliferating and 3 senescent monocyte samples, using NanoParticles or Neat (control). Only proteins present in at least 4 of 6 samples, and showing differential expression (Pval < 0.05, t-tests) in neat samples were included (n = 134) F) G) For all figures, human mapped peptides and proteins were used. **c,** CV (SD / Mean) was calculated for all proteins present in at least 2 of 3 Senescent samples and 2 of 3 Proliferating samples in Both Neat and NP MaxRep rollup.

### Supplemental Figure 3

**A) Comparing Correlation Coefficients of Linear Models using ENSPs + Controls, ENSPs only, and Control-Only Models**

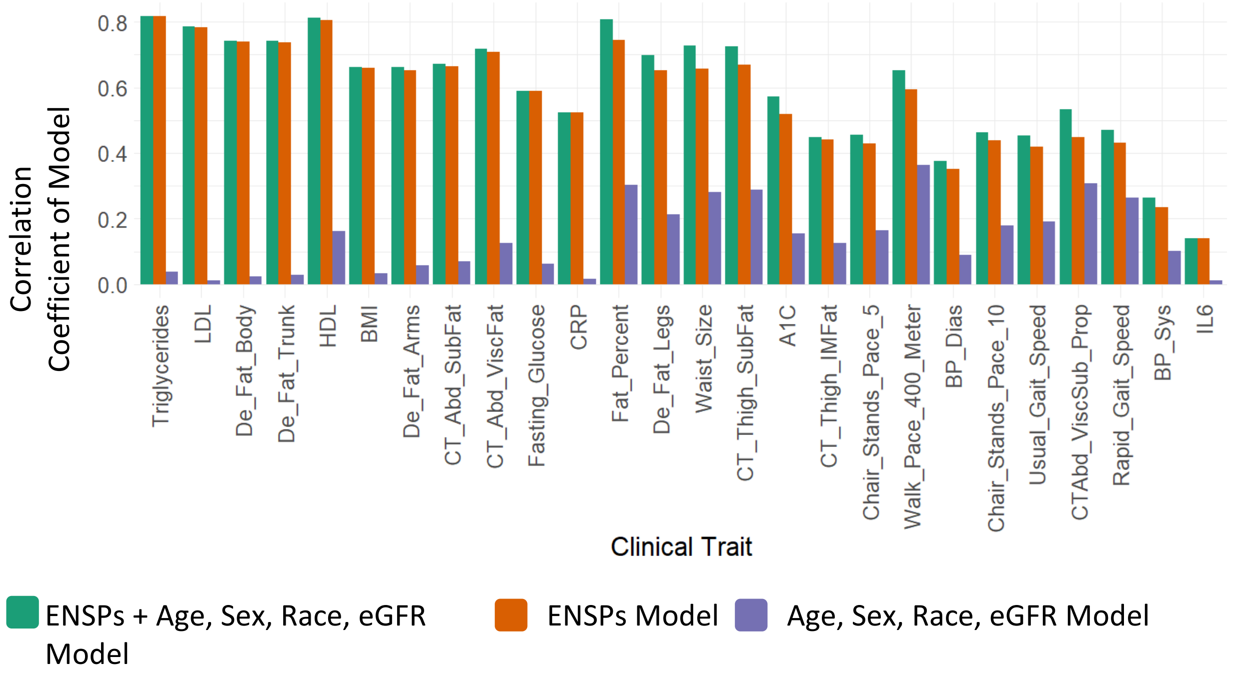


**B) Comparing Correlation Coefficients of Linear Models using ENSPs + Controls, ENSPs only, and Control-Only Models**

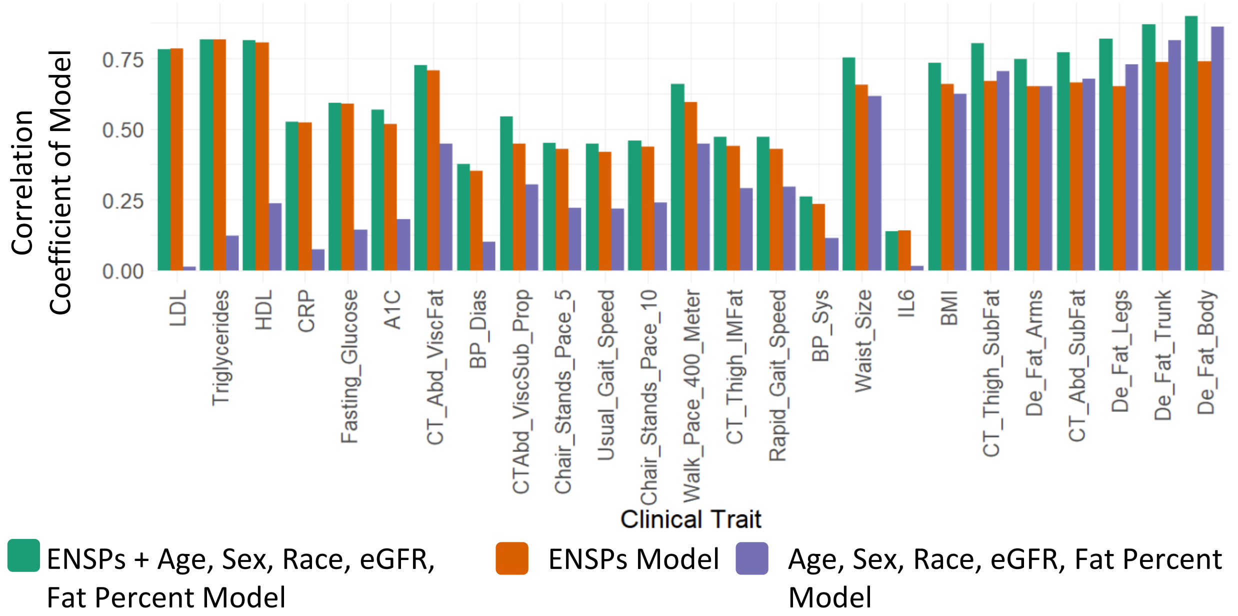


**Fig S3. ENSPs Show Age-Independent Predictive Potential. a,** Pearson linear models were constructed using covariates only (age, sex, race, and eGFR), ENSPs only, or ENSPs and covariates. Correlation coefficients for all models are shown by trait. **b,** This analysis is repeated but including fat percent as a covariate. Pearson linear models were constructed using covariates only (age, sex, race, and eGFR, and fat percent), ENSPs only, or ENSPs and covariates. Correlation coefficients for all models are shown by trait

### Supplemental Figure 4

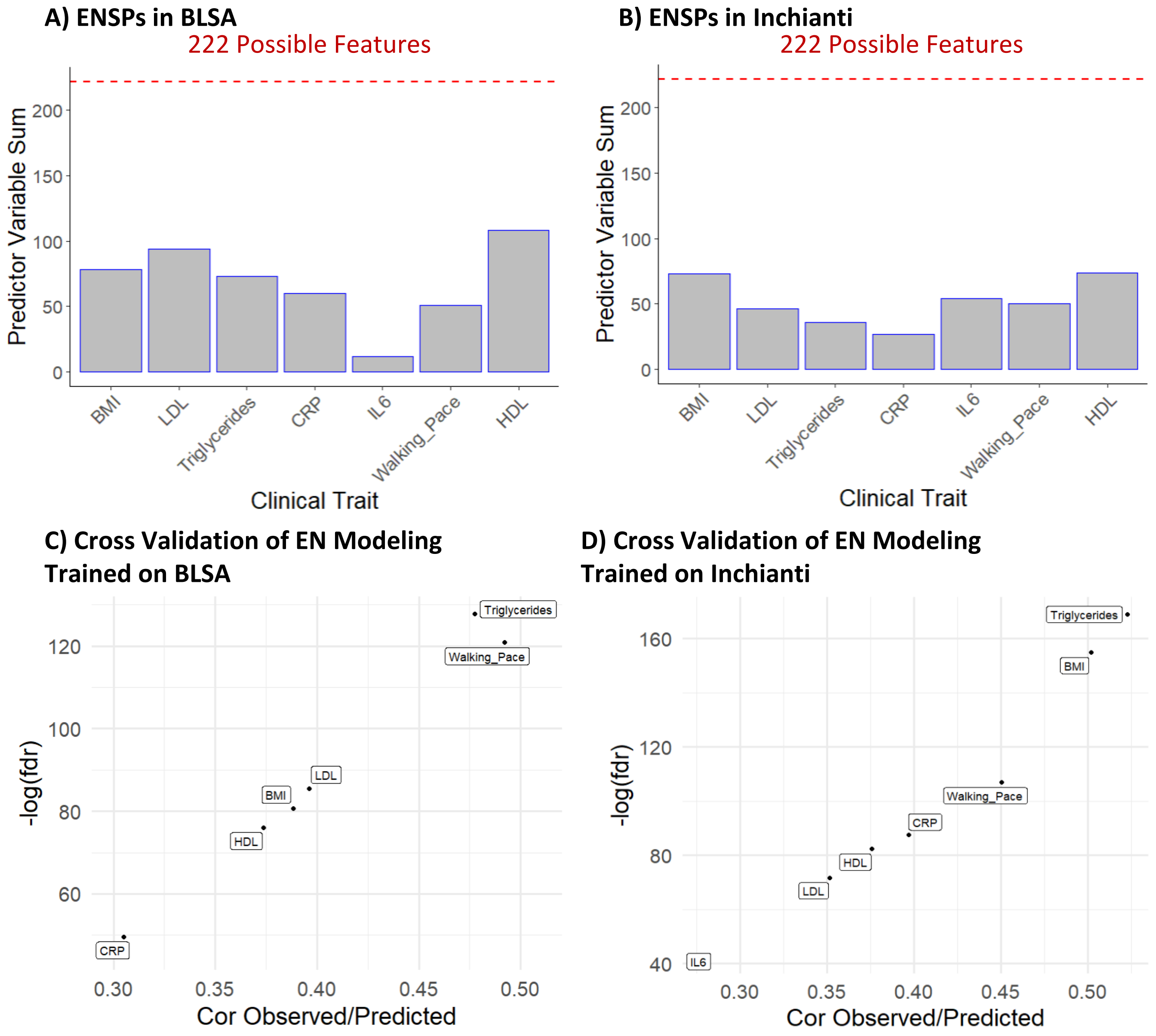


**Fig S4. Elastic Net Selected Proteins by Cohort**. **4 ,** 220 monocyte SASP were detected in both the BLSA (7k SomaScan) and Inchianti (1.3k SomaScan). Elastic Net modeling was used for feature selection in both Inchianti and BLSA. The number of ENSPs selected via elastic net for each train in the BLSA are shown. **b,** ENSPs selected in InCHIANTI. **c,** Linear models were constructed using only proteins selected in both studies for each trait. Spearman's correlation of predicted values of linear models trained on the BLSA and observed values in InCHIANTI. **d,** Spearman's correlation of predicted values of linear models trained on InCHIANTI and observed values in the BLSA.

### Supplemental Figure 5

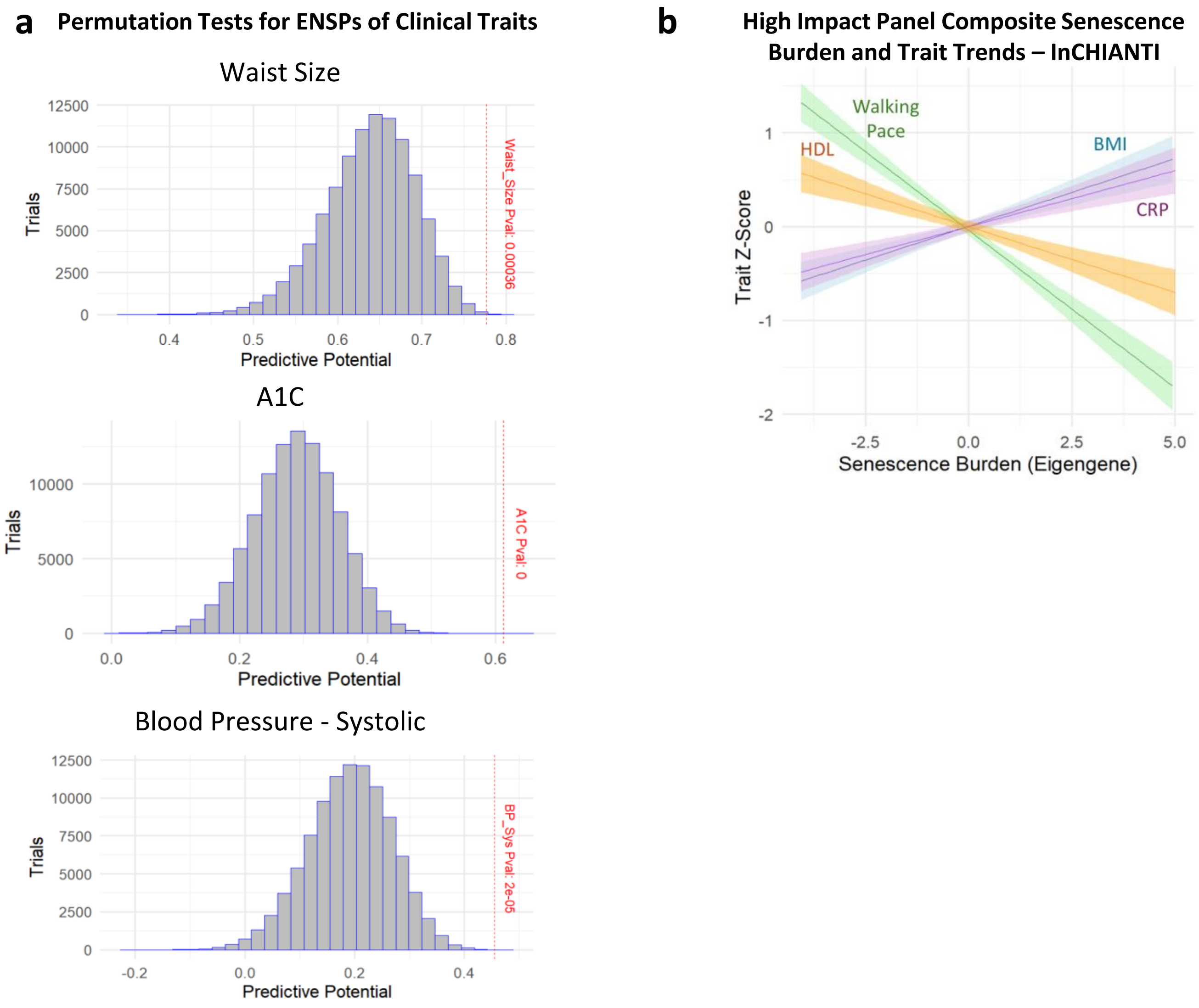


**Fig S5. Permutation tests and senescence eigengene in InCHIANTI**. **A)** Permutation tests comparing the predictive potential is shown for ENSPs compared with randomly selected proteins. Linear models for each trait were created either using ENSPs or randomly selected proteins of the same size. Models were trained on 80% of the data and used to predict the clinical traits for the remaining 20%. Randomly selected proteins models were trained and tested 100,000 times per trait and compared with the accuracy of the ENSP-only model. Red dotted lines show where the Spearman's correlation of the ENSP-only model lies in relation to the bell curve for the randomly selected protein models. **b,** Principal Component Analysis was used to condense the high-impact panel into a composite senescence burden score in the BLSA. Principal Component 1 was used to represent an eigengene for the high impact panel. With the InCHIANTI cohort ranked from low to moderate to high senescence burden, linear trait trends reveal that positive traits HDL and Walking Pace show a negative trend, while negative traits BMI and CRP show a positive trend.
